# Clinical Significance of a combination of Biochemical Indexes and Immunity Indexes for hepatitis B disease Prediction

**DOI:** 10.1101/2021.06.14.21258893

**Authors:** Wenyan Zhang, Shan Chen, Junzheng Yang

**Author notes:** Correspondence to Junzheng Yang, Quality Research Division, Guangzhou Packgene Co. Ltd, Guangzhou, Guangdong, 510000, China.

## Abstract

**Objectives:** To explore the clinical significance of the detection of biochemical indexes and immune indexes in the prediction of hepatitis B (HBV) disease.

**Methods:** 150 patients with HBV disease who were treated in our hospital from 2018 to 2020 were selected and divided into acute HBV group (group A), chronic HBV group (group B) and Cirrhosis group (C group), every group included 50 cases, and 50 healthy people were selected as the control group. Peripheral venous blood was collected from subjects from 4 groups, and immune indexes (plasma immunoglobulin G (IgG), plasma immunoglobulin A (IgA), plasma immunoglobulin M (IgM)) of HBV were measured by immune nephelometry, and biochemical indexes (alanine aminotransferase (ALT) and amino acid transaminase (AST)) of HBV were measured by fasting rate method.

**Results:** Compared with the control group, the immune nephelometry results and fasting rate method showed that the expression of immune indexes (IgG, IgA and IgM) and the expression of biochemical indexes (ALT and AST)in group A, B and C patients were increased significantly (P <0.05), and both of the expression of immune indexes and the biochemical indexes in HBV patients showed an increasing trend accompanied with the aggravation of HBV patients (A<B <C) (P<0.05).

**Conclusion:** The detection of immune indexes and biochemical indexes has important clinical significance in prediction of HBV patients’ condition, and detect a combination of biochemical indexes and immune indexes can be used to make a comprehensive judgment to provide a reliable laboratory basis for diagnosis of HBV patients’ condition.

## 1. Introduction

HBV is growing a global public health problem, there are 112 million long-term carriers of HBV patients in China, and about 0.75 million people die of cirrhosis and liver cancer caused by HBV infection every year^[1]^; in the world, About 2 billion people have been infected with HBV virus, and about 400 million people are carriers of HBV^[2]^. The first goal of clinical treatment for HBV is to inhibit the replication of HBV virus, and the long-term goal is to ameliorate the clinical symptoms of patients, prevent cirrhosis and liver cancer. Early pre-diagnosis of HBV patients’ condition can provide basis for the formulation of later treatment, and also an important means to improve the prognosis of patients with HBV. In the past, the detection of liver biochemical indexes was used to judge the patients’ condition. In recent years, with the further research on HBV pathology, some papers showed that T-cell immune response could result in liver cell damage, immune regulation function and immune response disorder after patients are infected with HBV virus, Those evidences suggests that immune indexes may be applied for prediction for HBV patients’ condition^[3]^. For this purpose, we collected 150 patients with HBV disease who were treated in our hospital from 2018 to 2020, try to analyze the connection between the combination of biochemical indexes and immune indexes and prediction of the patients’ condition.

## 2 Data and Methods

### 2.1 General information

Methods: 150 HBV patients have been treated in our hospital from 2018 to 2020 were selected and divided into four groups: acute HBV group (group A), chronic HBV group (group B) and liver cirrhosis group (Group C) according to the patient’s condition, each group included 50 cases, and 50 healthy people were selected as the control group. The study was approved by the medical ethics committee of Hebi infectious disease hospital, and informed consent was signed by patients and their families. In group A, there were 26 males and 24 females; The age ranged from 40 to 70 years old, with an average of (55±47) years old; There were 23 cases of hypertension, 19 cases of diabetes and 10 cases of coronary heart disease. In group B, there were 27 males and 23 females; The age ranged from 40 to 70 years old (55.78± 03) years old; There were 20 cases of hypertension, 17 cases of diabetes and 9 cases of coronary heart disease; In group C, there were 23 males and 27 females; The age ranged from 40 to 70 years old, with an average of (56.07±12) years old; There were 22 cases of hypertension, 21 cases of diabetes and 11 cases of coronary heart disease. in the control group, there were 25 males and 25 females; the age ranged from 40 to 70 years old, with an average of (56.11±24) years old. There was no significant difference in the general information of the four groups (P >0.05)(table 1).

**Table1.** The general information of participants in four groups

| Group | Male | Female | Age | Average age | Hypertension | Diabetes | Coronary disease |
| --- | --- | --- | --- | --- | --- | --- | --- |
| Group A | 26 | 24 | 40-70 | $55\pm47$ | 23 | 19 | 10 |
| Group B | 27 | 23 | 40-70 | $55.78\pm03$ | 20 | 17 | 9 |
| Group C | 23 | 27 | 40-70 | $56.07\pm12$ | 22 | 21 | 11 |
| Control | 25 | 25 | 40-70 | $56.11\pm24$ | None | None | None |
| $P>0.05$ | | | | | | | |

#### Inclusion criteria

(1) A, B and C all met the diagnostic criteria of HBV in the expert consensus on the mode of HBV test and diagnosis report^[4]^, and the results were confirmed by detection of HBV virus core associated antigen; (2) Serum HBsAg was positive for more than 6 months; (2) HBV DNA load≥1×10^4^ IU/mL。

#### Exclusion criteria

(1) patients in pregnancy or lactation period; (2) A history of alcohol abuse; (3) Systemic antiviral and immunomodulatory therapy; (4) Incomplete clinical data; (5) Patients with parenchymal organ dysfunction; (6) History of liver surgery.

### 2.2 Method

5mL of peripheral venous blood was collected from all participants in the four groups. serum extracted by centrifuge at 4°C and stored at −80°C. IgG, IgA and IgM were measured by immune nephelometry, and ALT and AST were measured by fasting rate method; The reagent was purchased from Antu biotaikang Company, and the automatic biochemical analyzer was purchased from Toshiba tba-120.

### 2.3 Statistics

The data involved in this study were analyzed by software SPSS22.0, in which the count data was expressed as %, and Χ^2^ test and variance analysis were used. *P*<0.05 was considered statistical significance.

## 3 Results

### 3.1 comparison of biochemical index results

The biochemical indexes ALT and AST in group A, group B and C were higher than those in the control group (P<0.05), and the expression of ALT and AST increased with the aggravation of HBV (A<B<C). The results were shown at Table 1.

### 3.2 comparison of immune indexes

The expression of IgG, IgA and IgM in group A, group B and C were higher than those in the control group (P<0.05), and the expression of IgG, IgA and IgM showed an increasing trend with the aggravation of HBV (a<B<C) (P<0.05). The results were shown at Table 2.

**Table 2.** Compared the expression of biochemical indexes (ALT and AST) among four groups 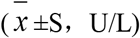

| among four groups ($\bar{x} \pm S$, U/L) | | |
| --- | --- | --- |
| Group | ALT | AST |
| Group A | 41.45±3.71 | 36.98±3.70 |
| Group B | 53.71±2.30 | 46.41±2.90 |
| Group C | 63.38±3.70 | 59.30±2.20 |
| Control | 26.62±2.74 | 23.65±3.24 |
| F | 2.708 | 3.656 |
| P | 0.021 | 0.032 |

**Table 3.** Compared the expression of immune indexes (ALT and AST) among four groups 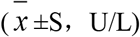

| among four groups ($\bar{x} \pm S$, U/L) | | | |
| --- | --- | --- | --- |
| Group | IgG (mg/dL) | IgA (mg/L) | IgM (mg/L) |
| Group A | 1535.38±54.60 | 2635.35±42.15 | 1984.65±62.23 |
| Group B | 1993.65±49.50 | 3961.11±19.60 | 4014.84±50.30 |
| Group C | 2695.52±51.23 | 4601.25±61.12 | 5967.68±52.25 |
| Control | 1319.25±24.74 | 2231.35±52.35 | 1756.71±43.47 |
| F | 2.473 | 3.135 | 2.401 |
| P | 0.028 | 0.021 | 0.039 |

## 4 Discussion

HBV is a liver disease caused by HBV virus infection. About 2 billion people in the world have been infected with HBV virus, and about 400 million people are carriers of HBV^[2]^. In most cases, HBV will not affect the normal life of patients, but HBV is an incurable disease, which is easy to progress into liver cirrhosis and liver cancer after long-term infection. In recent years, with the progress of medical science and technology, some scholars have given different treatment schemes according to different degrees of patients’ condition, the results showed that it has significant effect in preventing long-term complications and delaying the development of the disease^[5]^. Therefore, accurate identification and prediction of HBV is of great significance to improve the prognosis of patients. In the past, it was generally believed that the liver cell damage of HBV patients was caused by the replication of virus in the body, so the biochemical indexes of liver damage were often used to predict the patient’s condition. Now with the development of HBV research, T cell immunity plays an important role in the development of HBV. In view of this, the author selects immune indexes combined with biochemical indexes to predict the condition of patients with hepatitis B and discusses its clinical significance.

ALT and AST are the most commonly biochemical indexes in the diagnosis of hepatobiliary diseases. In patients with acute hepatitis, the activity of ALT in serum is increased, and the increase degree is higher than that of AST. To a certain extent, the increase range reflects the severity of liver cell damage in patients. When the patient’s condition improves, the activity also decreases. However, some studies reported that^[6]^ the level of HBeAg negative patients was lower than that of HBeAg positive patients, but the degree of liver injury was higher than that of HBeAg positive patients. This shows that although ALT and AST levels can effectively reflect the inflammation of patients with HBV, a certain detection value only reflects the liver injury of patients at a specific time point, and cannot fully reflect the degree of liver tissue injury of patients. At the same time, ALT and AST increased in patients with active HBV virus replication, suggesting that HBV virus can aggravate the inflammatory activity of patients in a short time, and the inflammation of patients’ liver tissue can simultaneously reflect the accumulation of short-term and long-term inflammatory activity. Therefore, ALT and AST levels may not be parallel to the patient’s condition. In this paper, the expression of biochemical indexes of ALT and AST in group A, B and C were higher than those in the control group (P<0.05), and the expression of ALT and AST showed an increasing trend with the aggravation of HBV (A< B<C) (^P^<0.05). We could explain the phenomenon as follows: (1) the course of disease in group A, B and C is A< B<C, and the accumulation degree of inflammatory injury in liver tissue is C>B>A, which leads to the increasing trend of ALT and AST with the aggravation of HBV; (2) HBV virus replication was in active phase and immunoglobulin was in high level in three groups.

It has been reported^[7]^ that after patients are infected with HBV virus, the damage of liver cells is not caused by HBV virus directly entering into liver cells for replication and reproduction, but by HBV virus entering into the body to stimulate a series of immune reactions in patients, in other word, HBV virus is a non-cytotoxic virus, which does not directly damage the body, but promotes the immune dysfunction of patients. After the body is infected with HBV virus, its antigen will continue to exist in the liver cells and tissues, stimulate the body to produce B cells, disorder of immune function quickly promote B cells into plasma cells, and produce specific antibodies, namely plasma immunoglobulin. At the same time, it has been reported^[8]^ that the development of chronic HBV is mainly due to the dysfunction of Kupffer cells in liver cells, which makes it difficult to remove the absorbed antigens from the intestine in time, and the body produces too many antibodies, leading to the increase of immunoglobulin. The increased plasma immunoglobulins were mainly IgG, IgA and IgM. In this study, IgG, IgA and IgM in group B and C were higher than those in the control group (P<0.05), and the above indexes showed an increasing trend with the aggravation of hepatitis B (A< B<C) (P<0.05). The results showed that there were immune damage in patients with different degrees of HBV, and the immunoglobulin increased with the progress of the disease. The analysis shows that: humoral immunity in the body of patients with chronic HBV is in a state of hyperfunction, and humoral immunity is the main link in the process of HBV virus infection, body defense and the progress of HBV, so the plasma immunoglobulin will increase significantly after the body is infected with HBV. The abnormal increase of immunoglobulin level will directly lead to the decrease of the ability to dissolve virus antigen and immune complex, form a vicious circle, and aggravate the degree of liver cell immune damage. In addition, with the development of the disease, liver cell necrosis has occurred in patients with liver cirrhosis. The necrotic liver cells and tissues will aggravate the stimulation of the immune function of patients. The immune system will be more hyperactive and secrete antibodies to remove necrotic cells and tissues, resulting in higher levels of immunoglobulin in patients than those in groups A and B. Therefore, immunoglobulin can be regarded as an important indicator to predict the condition of patients with HBV.

In conclusion, the detection of biochemical indexes and immune indexes has important clinical significance in predicting the condition of HBV patients. In the clinical prediction of HBV, biochemical indexes and immune indexes can be combined for comprehensive judgment to provide reliable laboratory basis for HBV patients.

## Data Availability

The data used to support the findings of this study are available from the corresponding author upon request

## Acknowledgement

None.

